# Mortality prediction by a metabolomics score and health- and lifestyle-related factors combined

**DOI:** 10.64898/2026.02.01.26345306

**Authors:** Kerstin Schorr, Mar Rodriguez-Girondo, Lisette CPMG de Groot, P. Eline Slagboom, Marian Beekman

## Abstract

The ageing society and worldwide rise of chronic disease make adequate early identification of at-risk individuals and preventive intervention highly relevant to public health. Molecular indicators of global health have been developed, such as metabolomics-based MetaboHealth. A shortcoming of molecular biomarkers may be their lack of integration of lifestyle and environmental factors relevant for health span. Hence, we explored the MetaboHealth biomarker and a range of health- and lifestyle factors, including plant based diet index, physical activity, alcohol use, smoking, medication use, 25(OH)D status and socioeconomic position and education in a subpopulation (n=35,192, mean age=56 years) from the UK Biobank cohort. We analysed which of these factors associated independently with mortality; which associated with the MetaboHealth score and which of the independent factors improve mortality prediction by MetaboHealth. By applying multivariate Cox regression modelling we found that all factors associated independently with prospective survival, except for physical activity and education level. Sex, smoking and income were most strongly associated with both mortality and the MetaboHealth score. By cross-validation we subsequently assessed contribution of all independent health- and lifestyle-related factors to MetaboHealth-based mortality prediction and computed a weighted score. We found income and medication intake to be the most and diet the least prominently adding contributors. In conclusion, MetaboHealth partly reflects the effect of health- and lifestyle-related factors, while identification of at-risk individuals is improved by the information on income and medication use. Insights in these factors can be attained non-intrusively and may therefore be taken into account in the context of population health management.

## 1. Introduction

The ageing population and the concomitant rise in chronic diseases have been widely discussed as a major societal challenge (1). Large inter-individual heterogeneity exists in the individual rate and nature of ageing (2). Adequate identification of individuals at risk for chronic disease is crucial to direct resources and prevention strategies towards vulnerable populations and therefore is ultimately beneficial for public health. Knowledge about the contribution of separate lifestyle and environmental factors is relevant for care providers to be able to spot at-risk groups in population health management. To establish such risk, predictors have been generated based on combinations of risk factors. The most commonly known is the Framingham Risk Score (FRS), developed to predict individual risk of developing heart disease over a 10 year time frame by combining age, sex, HDL and LDL cholesterol levels, blood pressure, and smoking in an algorithm. The FRS has been used to identify at-risk individuals and helps in determining treatment strategies (3, 4). However, the FRS is focused specifically on coronary heart disease (CHD), leaving out other aspects crucial to the global health of ageing individuals. Additionally, it has been found to perform less well for younger men and leaves out aspects such as family history of cardiovascular disease.(5, 6). Further, while the FRS does take into account smoking, other lifestyle factors relevant to health are not considered. Therefore, to study individuals at risk of accelerated ageing and decline of global health, the FRS is not sufficiently informative.

In recent years, interest has grown in generating molecular blood based markers indicative of the decline of global health in older adults. Such algorithms generated from a.o. epigenetic, proteome and metabolome data, were focused on predicting chronological age (and the acceleration thereof), ageing phenotypes and clinical endpoints, risk factors and mortality. One of the predictors trained to predict mortality across all ages is the MetaboHealth score, a metabolomics based algorithm consisting of 14 circulating metabolite markers (7). MetaboHealth was trained in one of the largest studies of its kind and has shown to represent various aspects of global health, especially in older individuals. The score is predictive of cognitive decline and frailty, while outperforming chronological age-trained biomarkers (8-10). Since MetaboHealth has potential to be used as global health indicator in lifestyle improvement strategies in population management or primary care, the question raises which health and lifestyle factors are captured by MetaboHealth and how its power to predict mortality and global health decline may be improved. Some of the relevant factors are well known and have been studied extensively in the past, including age, sex, body mass index (BMI), alcohol and smoking (11). Apart from that, a plant-based diet, has been found to be associated with mortality risk as well as with markers of metabolic health in previous research, potentially due to antioxidative and anti-inflammatory effects of plant foods (12-14). Research also points out the role of 25-hydroxyvitamin D (25(OH)D) and physical activity in decreasing mortality risk in older adults (15, 16). 25(OH)D has been shown to regulate biological pathways, thereby potentially contributing to decreased cancer risk (17). Another factor associated with mortality risk in older adults is polypharmacy (18). Lastly, lower socio-economic status (SES) has been identified as a factor associated with an unfavorable metabolic profile and it is known that low income is associated with shorter health- and life span (19-21).

The most promising efforts to build informative algorithms indicative of global health (or biological age) by focusing on mortality have not included lifestyle factors. Indeed little attention has been paid to generate health scores based on lifestyle and molecular data combined. Generating health-scores combining molecular, lifestyle and socioeconomic position may have the potential of better identification of individuals at risk for age-related disease.

Here we assess which of a set of health-and lifestyle-related factors are associated with prospective mortality, independent of the established metabolomics-based biomarker MetaboHealth. This set includes age, sex BMI, plant based diet index, physical activity, alcohol use, smoking, medication use, 25(OH)D status and socioeconomic position and education. Secondly we investigate which of these factors associate with MetaboHealth to explore which information in these factors is represented by the molecular score. Lastly, we investigate if addition of the separate health-and lifestyle related factors improves the prediction of all-cause mortality in addition to MetaboHealth and include these in a joint model to generate a better biomarker, incorporating metabolic and lifestyle data, for global health able to identify persons at risk for age-related disease.

## 2. Methods

### 2.1. Study Population

For this study, we used data from the UK Biobank, a multicenter study with >500,000 participants conducted in England, Scotland and Wales. Ethical approval was granted by the North West Multi-centre Research Ethics Committee and all participants provided written consent at recruitment (22). Baseline measurements were carried out between 2006 and 2010. For the purpose of this study, we included participants that had baseline data on all variables of interest available and filled at least two 24h-diet recalls (Suppl. Table 1). Our final sample consisted of n=35,192 (mean age=55.5 years) (Table 1).

**Table 1.** characteristics in joint sample and by sex.

|  | **Joint** | **Female** | **Male** |
| --- | --- | --- | --- |
| **N** | 35,192 | 18,270 | 16,922 |
| **Age in years** | 55.5±7.8 | 54.7±7.7 | 56.5±7.9 |
| **Female n%** | 18,270 (51.9%) |  |  |
| **hPDI, mean ± SD** | 53.2±7.5 | 53.1±7.5 | 53.3±7.4 |
| **uPDI, mean ± SD** | 52.7±7.3 | 52.2±7.3 | 53.3±7.2 |
| **MetaboHealth, mean ± SD** | -0.09±0.43 | 0.02±0.41 | -0.21±0.43 |
| **Income, n%** |  |  |  |
| **Low** | 4,167 (11.8%) | 2,463 (13.5%) | 1,704 (10.1%) |
| **Medium** | 18,486 (52.5%) | 9,726 (53.2%) | 8,760 (52.8%) |
| **High** | 12,539 (35.6%) | 6,081 (33.3%) | 6,458 (38.2%) |
| **Education, n%** |  |  |  |
| **Low** | 8,089 (23.0%) | 4,494 (24.6%) | 3,595 (21.2%) |
| **Medium** | 6,801 (19.3%) | 3,278 (17.9%) | 3,523 (20.8%) |
| **High** | 20,302 (57.7%) | 10,498 (57.5%) | 9,804 (57.9%) |
| **BMI [kg/m<sup>2</sup>], mean ± SD** | 26.6±4.4 | 26.1±4.8 | 27.1±4.0 |
| **Physical Activity (h/wk), mean ± SD** | 39.1±37.1 | 38.6±35.9 | 39.6±38.3 |
| **Nr of medication, mean ± SD** | 2.0±2.3 | 2.1±2.3 | 2.0±2.4 |
| **Vitamin D [nmol/l], mean ± SD** | 49.6±20.8 | 49.3±20.6 | 49.9±21.0 |
| **Smoking (Current), n%** | 2360 (6.7%) | 1,036 (5.7%) | 1,324 (7.8%) |

hPDI=healthful plant-based diet index, uPDI=unhealthful plant-based diet index, BMI= body mass index, income categories: low (<18,000£), medium (18,000 – 51,999£), and high (>52,000£)

### 2.2. Plant-based diet index

For the calculation of the plant-based diet index, we used preprocessed data that provided information on dietary intakes of 93 food groups in grams per day (Category 10018, Suppl. Table 1). This dataset was created by Piernas et al. in 2022 based on raw data from the Oxford WebQ tool by linking it to food composition data (23, 24). In this web-based, self-administered questionnaire, participants reported their intake in serving sizes within the last 24h. Invitations to complete the questionnaire were sent on specific days to capture variations in intake between weekdays and the weekend^1^. In our study, participants that completed at least two questionnaires were included in the analysis, to gain an estimate of usual intake. We further excluded participants with unrealistic energy intake as described previously (25). To calculate the PDI for each participant from this data set, intakes of 17 food groups, consisting of healthful plant foods (wholegrains, fruits, vegetables, nuts, legumes, tea & coffee), unhealthful plant foods (sweets, refined grains, fruit juices, sodas, potatoes) and animal foods (meat, fish, dairy, eggs, animal fats, miscellaneous animal foods), were averaged across questionnaires (Suppl. Table 2). Vegetable oil was not included since insufficient information was available. The PDI was calculated following a standard procedure (26): The intakes per food groups were divided into sex-specific quintiles. For the healthful plant-based diet index (hPDI) this includes scoring healthful plant foods positively, whereas unhealthful plant foods were scored negatively. For the unhealthful plant-based diet index (uPDI) this procedure was reversed: healthful plant foods were scored negatively and unhealthful plant foods were scored positively. Animal food groups were always scored negatively for both indices. Since intakes in some food groups were low, the cut-off for several quintiles was 0. Therefore, quintiles with a low intake (0 g until the cut-off of the first non-zero quintile) were summarized and awarded a 1. The remaining quintiles were then scored as normal, i.e., those above the fifth quintile receiving a score of 5 etc. The final score has a theoretical score ranging from 17-85, with higher scores indicating greater adherence to a healthful (hPDI) or unhealthful (uPDI) diet, respectively. The index was adjusted for energy intake using the residual method (27). For this analysis, those in the top tertile of the hPDI or uPDI are compared to the rest of the study population, in order to estimate effect size among those with greatest adherence to the respective dietary pattern.

### 2.3. Assessment of metabolic Health

MetaboHealth is a marker consisting of 14 metabolites independently associated with mortality, selected out of 65 markers measured by the Nightingale platform (7). The MetaboHealth score has been derived from over 44 000 individuals out of 12 cohort studies (not including the UK Biobank), spanning a wide age-range, by applying a forward-backward approach. MetaboHealth can be calculated via a code available on the Github of D. Bizzari (https://github.com/DanieleBizzarri/MiMIR).

The 14 metabolites include : total lipids in chylomicrons and extremely large very low-density lipoprotein (VLDL) particle, total lipids in small high-density lipoprotein, mean diameter for VLDL particles, ratio of polyunsaturated fatty acids to total fatty acids, glucose, lactate, histidine, isoleucine, leucine, valine, phenylalanine, acetoacetate, albumin, and glycoprotein acetyls. MetaboHealth ranges from -2 to 3 in most cohorts and a 1-unit increase in MetaboHealth has been associated with a 2.73 times higher mortality risk (7). A higher score therefore represents higher mortality risk. For this analysis, we compare those in the top tertile of MetaboHealth to the rest of the study population, in order to estimate effect sizes among the most at-risk population..

### 2.4. Outcome assessment

In the UK Biobank, mortality data is derived from national death registries. Censoring dates for death data were provided by the NHS Information Centre for participants from England and Wales until November 2023, and the NHS Central Register Scotland for Scotland until December 2023. Time in the study was used as the time scale in all analyses, and the time-to-event variable was defined as the difference between the age at last follow-up or event occurrence, whichever occurred first, and the age at baseline In total 1782 deaths were observed. On average, participants were followed for 15.3±1.8 years from baseline to censoring date. To model time to death, we used Cox proportional hazards regression.

### 2.5. Health- and lifestyle related factors

As health- and lifestyle-related factors, due to their known association with mortality, we considered age, sex, BMI (28, 29), adherence to an (un)healthful plant-based diet (12, 14), smoking and alcohol intake (in g/d) (11), number of prescriptions or over-the-counter medication (18), amount of physical activity (in MET h/wk) (16), education level (21), income (20) and 25(OH)D (nmol/l) (17). All models are further adjusted for UKB assessment center to account for potential differences in data collection between centers. Total household income before tax was coded into three categories: low (<18,000), medium (18,000 – 51,999), and high (>52,000). Education was categorized into low (CSEs or equivalent, O levels/GCSEs or equivalent), medium (A levels/AS levels or equivalent, NVQ or HND or HNC or equivalent) and high (College or University degree, Other professional qualifications e.g.: nursing, teaching).

### 2.6. Statistical Analysis

#### 2.6.1. Multivariate Cox regression modelling

We first assessed which health- and lifestyle related factors are associated with mortality risk independent of each other and MetaboHealth, in order to select which variables to include in the following step of building a predictor. To this end, a Cox proportional hazards model was fitted including all these factors. Because the diet variables hPDI and uPDI are correlated, we fit separate models for each diet index, resulting in Model 1.1 with hPDI and 1.2 with uPDI.

Model 1.1: Time to mortality ∼ MetaboHealth + hPDI + health-&lifestyle covariates

Model 1.2: Time to mortality ∼ MetaboHealth + uPDI + health-&lifestyle covariates

Secondly, we assess which health- and lifestyle-related factors independently associated with the MetaboHealth score via logistic regression analysis with binarized MetaboHealth (top tertile vs rest, cut-off=0.076) score as outcome. This allowed us to gauge how much variation of health- and lifestyle related factors is already captured by MetaboHealth. Because the diet variables hPDI and uPDI are correlated, we fit separate models for each diet index, resulting in Model 2.1 with hPDI and 2.2 with uPDI.

Model 2.1: MetaboHealth ∼ hPDI + health-&lifestyle covariates + sex

Model 2.2: MetaboHealth ∼ uPDI + health-&lifestyle covariates

#### 2.6.2. Assessing predictive performance

To further answer the question of the individual contribution of relevant lifestyle- and health-related variables to mortality prediction, we performed comparisons of model performances (c-indexes) of consecutive mortality prediction models:

Model a: Time to mortality ∼ age+sex+bmi+alcohol+smoking

Model b: Model a + metaboHealth

Model c: Model a + hPDI

Model d: Model a + uPDI

Model e: Model a+ Medication intake

Model f: Model a + income

Model g: Model a + vitamin D

Model h: Model a + metaboHealth + income + uPDI + vitamin D + medication intake

Model a is the basic model consisting of known contributors to mortality risk, namely age, sex, BMI, smoking and alcohol intake. Secondly, we added variables that were statistically significantly (p-value<0.05) associated with mortality in the previous step (Model 1.1 and 1.2, Table 2) separately to the basic model. In the final step, we add all predictors together to determine the predictive power of all variables combined. As previous research has found the MetaboHealth to be predictive particularly among older adults (7), we repeat prediction models in strata of age (<55 years and >=55 years) and in addition also in sex strata.

**Table 2.**
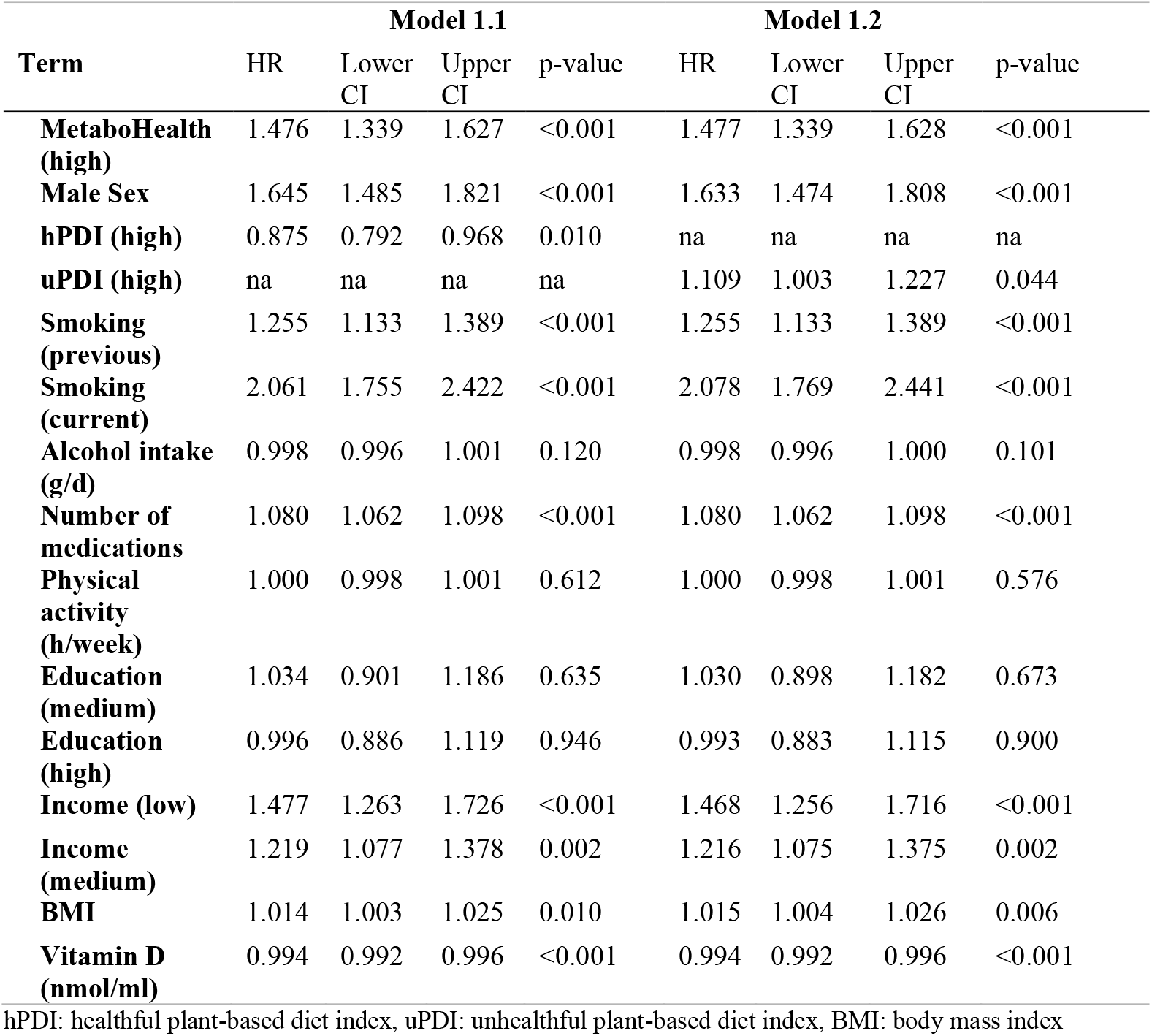
Associations of Health- and Lifestyle related factors with Mortality independent of MetaboHealth.

To validate our prediction models, we perform a leave-one region-out cross-validation procedure, using assessment centers grouped according to the geographical regions of the UK, meaning iteratively each region is left out of and used as test set (Suppl. Table 3), while the remaining data serves as training data. Model performance was assessed by calculating the concordance-index (c-index) (30). To compare the predictive performance of the models, we compare the c-index for each model. A higher c-index denotes better predictive power. The final prediction model was selected based on the highest c-index and its calibration was assessed using a calibration plot with a 15-year time horizon. A weighted score was then constructed by multiplying the model coefficients with the corresponding predictor values for each individual and summing the results.

## 3. Results

### 3.1. Characteristics of Study population

Our sample consisted of 35,192 participants with a mean age of 55.5±7.8 and contained slightly more women than men (52%). Women in our sample had slightly lower adherence to both hPDI and uPDI, were less likely to be smokers and had a slightly lower BMI. Men were somewhat less likely to be in the low income category (Table 1).

### 3.2. Health- and lifestyle factors independently associated with all-cause mortality

To first assess whether health- and lifestyle-related factors associate with all-cause mortality independently of each other and MetaboHealth and to make a selection for a subsequent prediction model, we added all health- and lifestyle related factors together with MetaboHealth in a multivariate model (Table 2). Besides MetaboHealth (HR=1.48 [95% CI 1.34, 1.63]), male sex, drug use and diet significantly associated with prospective survival. Current smoking showed the strongest association with all-cause mortality (HR=2.06 [95% CI 1.75, 2.42]), followed by male sex (HR=1.64 [95% CI 1.49, 1.82]) and the number of medication (HR=1.08 [95% CI 1.06, 1.10]. Belonging to the top tertile of adherence of hPDI and uPDI respectively (HR_I_=0.88 [95% CI 0.79, 0.97], HR=1.11 [95% CI 1.01, 1.23]) are both associated with prospective mortality. Additionally, the low income group had 48% higher mortality risk of mortality compared to high earners (HR=1.48 [95% CI 1.26, 1.73]). Lastly, VitD was associated with a reduced mortality risk (HR=0.99 [95% CI 0.99, 1.00]). Remarkably, physical activity and education level are not associated with prospective survival independently of the other factors.

### 3.3. Health- and lifestyle related factors associated with lower MetaboHealth score

To investigate whether MetaboHealth is partially correlated with the health- and lifestyle related factors, and thereby partially explains the association with mortality, we performed models 2.1 and 2.2. Significantly associated with MetaboHealth are hPDI, uPDI, age, BMI, physical activity, medication intake, income, sex, and smoking, the latter three having the largest effect size on MetaboHealth. The effect of male sex, plant-based diet adherence and income are partially captured by MetaboHealth, since these variables are associated with both, MetaboHealth and mortality. Meanwhile, the effect of physical activity on mortality is captured fully by MetaboHealth, as we observe an association with MetaboHealth but not mortality. The effect of vitamin D on mortality seems to be fully independent of MetaboHealth, since we observed a significant association with mortality but not MetaboHealth. (Suppl. Table 4).

### 3.4. Health- and lifestyle factors contribute to mortality prediction

Next, we explored the individual contribution to mortality prediction of health- and lifestyle factors independent of traditional mortality risk factors age, sex, bmi, alcohol and smoking. In order to assess the contribution of lifestyle factors to mortality prediction, we added our variables of interest separately to the basic prediction model and compared their respective c-index (Table 3, Model a). The contribution of each variable to the prediction are displayed in Table 3. The most influential variables in the prediction of mortality were MetaboHealth, medication intake and income, whereas hPDI and uPDI played only a small role (Table 3). Time to mortality was best predicted by MetaboHealth, medication intake, income, vitamin D and uPDI (C-index model h=0.739). Figure 1 shows the importance of variables in model h on mortality risk (Suppl. Table 5). According to the calibration plot at 15 years, the model showed reasonable calibration, with good agreement between observed and predicted survival probabilities (Suppl. Figure 1).

**Table 3.**
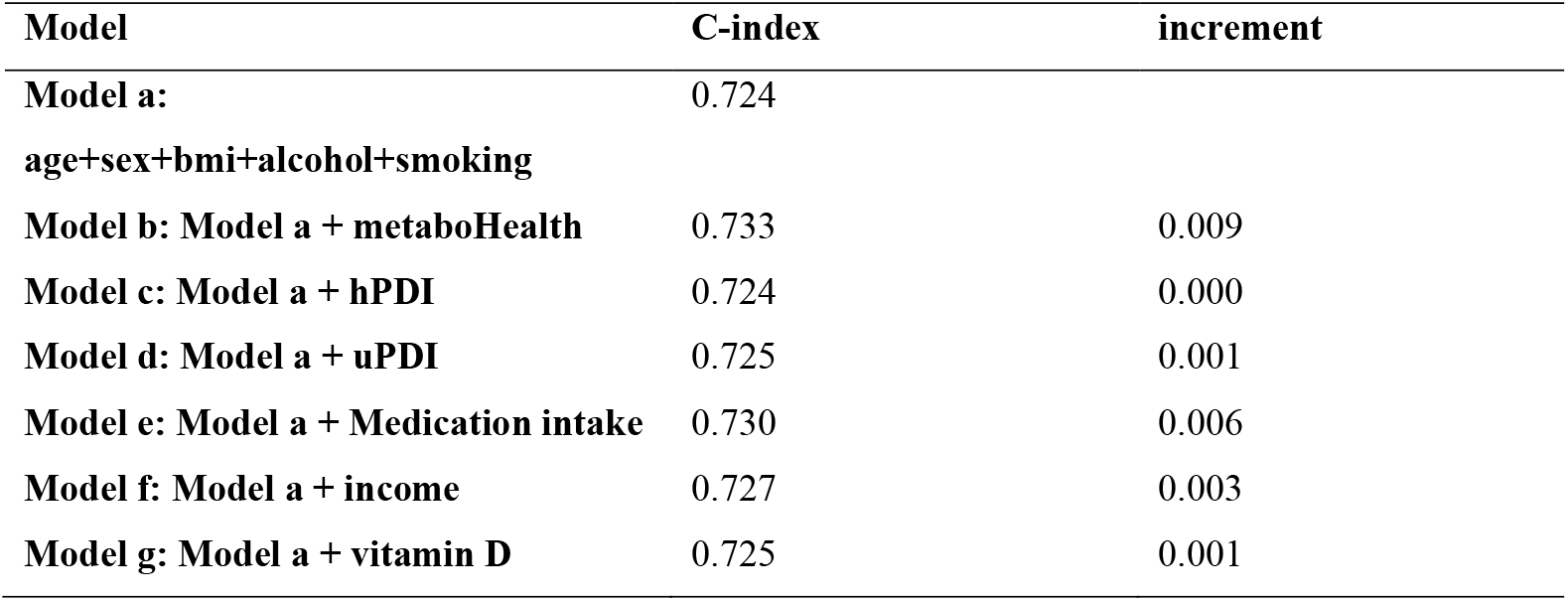
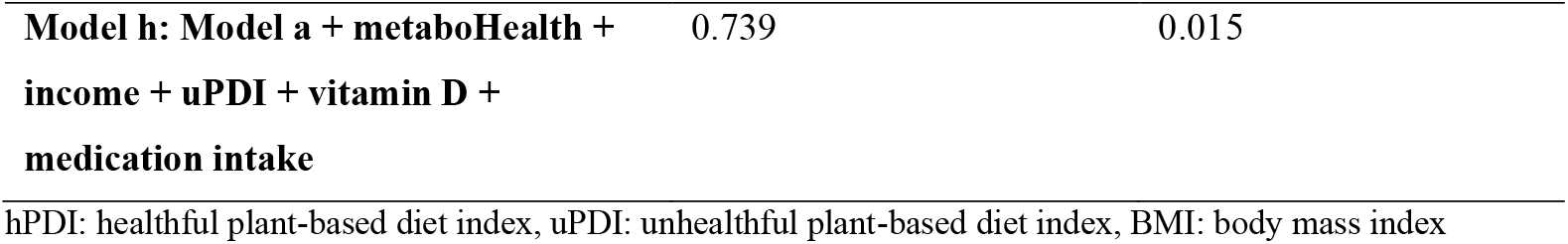
Overview over prediction models and individual contribution of health- and lifestyle factors to mortality prediction.

**Figure 1.**
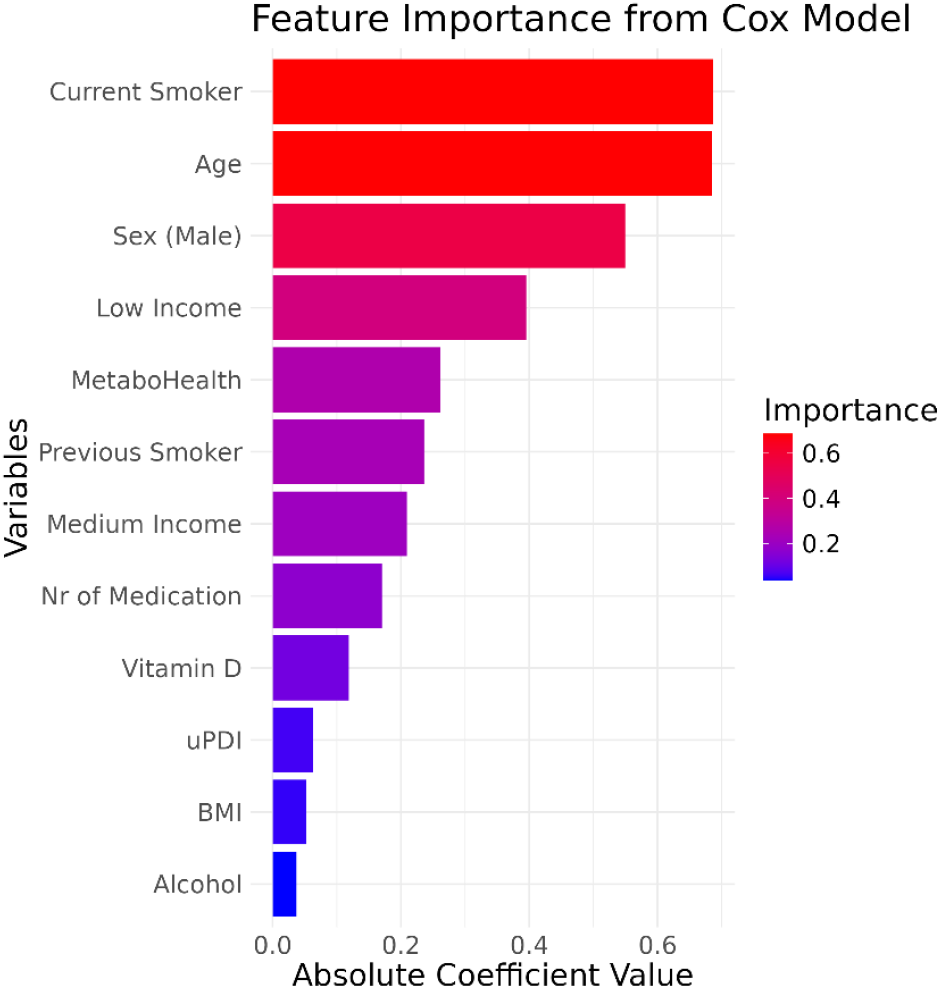
Feature importance plot showing individual contribution to mortality risk of each variable in the model (scaled) uPDI: unhealthful plant-based diet index, BMI: body mass index Created in R 4.4.1

Next, we computed a weighted score using the coefficients of predictors included in model h. The different survival curves of those with low and high mortality risk score (comparing the top tertile to the rest) are shown in Figure 2. Those in the high risk group were approximately 10 years older and had a higher MetaboHealth score (62.9±4.1, 0.08±0.4) compared to the low risk groups (51.9±6.6, - 0.17±0.4). Further, the high risk group had a higher BMI (27.8±4.6 vs 26.0±4.2 kg/m^2^ and was more likely to be male (Suppl. Table 6).

**Figure 2.**
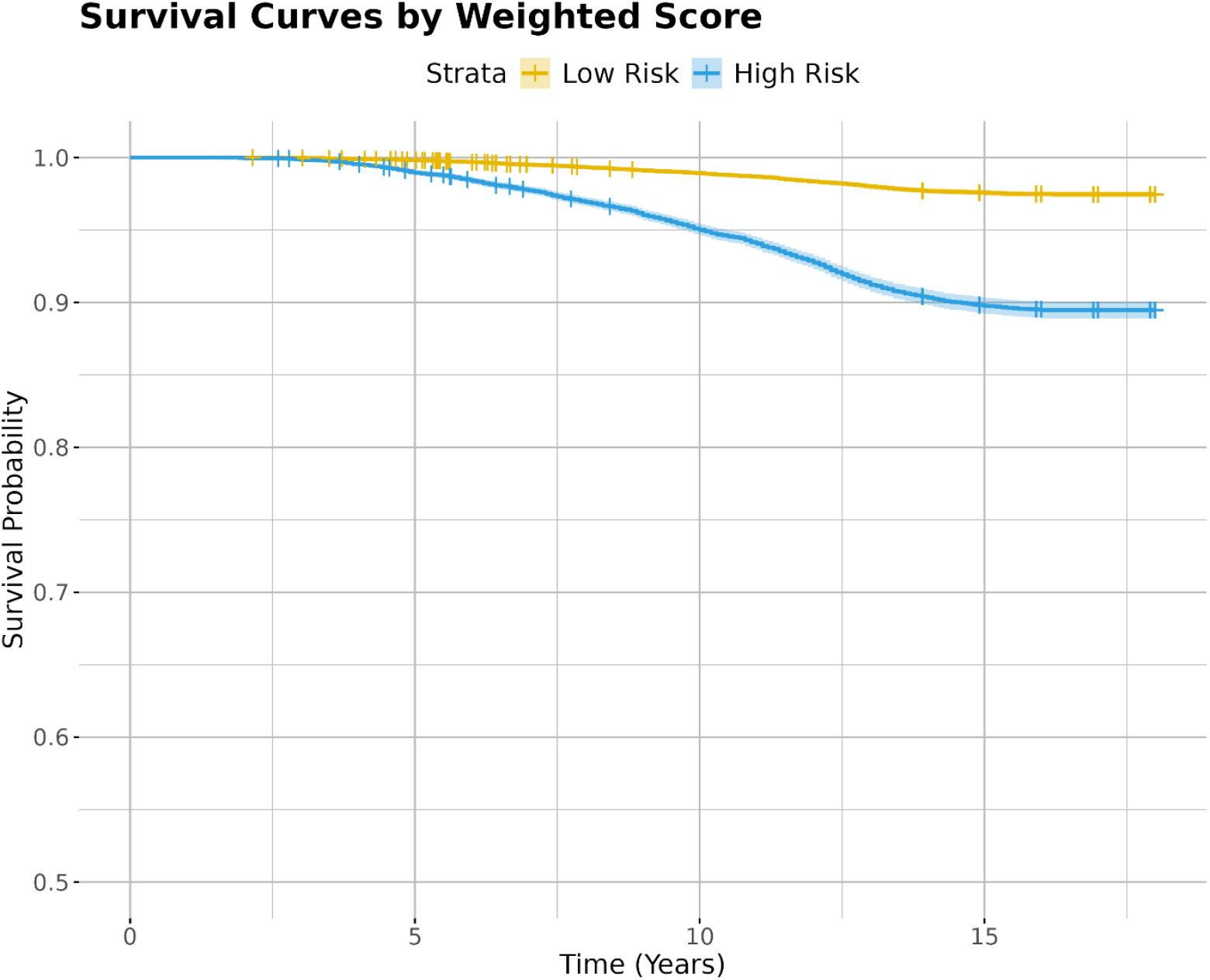
Survival curve for individual with low and high weighted risk scores (highest tertile vs rest) Created in R 4.4.1

To assess if there is a difference in the performance of our prediction model depending on sex or age, we repeated the prediction modelling process separately in younger (<55 years) and older adults (>=55 years) as well as stratified for men and women (Suppl. Table 7). We found that all determinants were slightly better at predicting mortality in older adults and in men. The full model h predicted mortality better than that based on traditional variables (Suppl. Table 7) most prominently in men. Further, medication intake was a stronger predictor in older adults compared to younger adults. Income is more predictive for mortality among men than among women. Diet had a marginal influence in all subgroups (Suppl. Table7).

## 4. Discussion

We investigated which health-and lifestyle-related factors independently associate with prospective mortality, independent of the molecular marker of ageing and global health MetaboHealth score (7). In addition, we built a predictor for mortality using all independent factors associating with mortality that we investigated. Our results show that smoking, medication intake, income, vitamin D and unhealthful plant-based diet adherence independently associated with mortality. While the prediction of mortality by MetaboHealth can partially be explained by correlation with adherence to an unhealthful plant-based diet, low income, male sex and high BMI, these health- and lifestyle related factors also independently contributed to the prediction of mortality. We built a global mortality predictor, in which currently being a smoker, male sex, having a low income and high MetaboHealth score showed the largest importance. Mortality could be better predicted in older adults (>=55 years) and in men. Taking into account income, medication intake and MetaboHealth, besides the well-established traditional risk factors age, sex, BMI, alcohol intake and smoking behavior, will improve the identification of especially older individuals at risk for disease in population health management.

MetaboHealth is a metabolomics-based marker of global immune-metabolic health, given it has been shown to associate with a variety of health issues like frailty, cognitive decline, and fracture risk (8-10, 31). Our results showed that MetaboHealth completely captured the effect of physical activity, and partially the effect of diet on mortality. In line with other research, we found a healthful plant-based diet associated with lower mortality risk and an unhealthful plant-based diet associated with higher mortality risk (12, 25, 32). Here, we showed that low MetaboHealth, which is indicative of low mortality and disease risk, is associated with a healthful plant-based diet. Meanwhile high MetaboHealth, which is indicative for high mortality and disease risk, is associated with adherence to an unhealthful plant-based diet. Probably due to partial correlation between PDI and MetaboHealth, we observed in our prediction models little to no contribution of diet on the predictive power. Remarkably, while low income was also associated with both MetaboHealth and higher mortality risk, low income had larger independent predictive value for mortality. In addition to the well-established determinants, MetaboHealth is the strongest novel predictor for mortality risk in our prediction model, showing its potential to discriminate low and high risk individuals. Overall, in association with mortality, MetaboHealth as a marker of global immune-metabolic health reflects physical activity completely and partially components of a healthy diet, non-smoking, high income, and low number of medications.

Our finding of hPDI being associated with lower MetaboHealth is novel, albeit not unexpected. Previous research has revealed beneficial effects of a healthful plant-based diet on circulating levels of blood lipids, amino acids and inflammatory markers individually, all of which make up the MetaboHealth (33-35). However, this analysis is the first to show that a healthful plant-based diet associates with a healthier immuno-metabolic profile predictive of mortality, frailty and cognitive decline. Along the same lines, the opposite finding for an unhealthful plant-based diet can be explained, as uPDI has been found to be associated with an adverse metabolic profile (33-35). Similarly, low SES has been shown to be associated with lower levels of HDL and higher circulating C-reactive protein, a marker for inflammation (20, 36). Accordingly, an association of low income with high MetaboHealth falls in line with previous findings. Health behaviors have been identified as contributors to the socioeconomic differences in health (37). However, in our analysis we found income to be associated with both, immune-metabolic and mortality risk after adjusting for behaviors such as smoking, alcohol intake and diet. This may suggest either the presence of residual confounding, e.g., through factors associated with SES such as poor housing or exposure to pollution or psychological distress or inadequate access to medical care (36, 38).

Current smoking most strongly associated with mortality risk in multivariate models, in line with previous findings strongly associating smoking with mortality risk. Interestingly, research has also revealed smoking to be a large contributor to socioeconomic inequalities in mortality. Since we also identified income as a relevant predictor for mortality, the overlap of smoking and income group may allow adequate identification of at risk groups. Lastly, higher number of medication was significantly associated with mortality risk and had the second largest contribution to our prediction model, which is unsurprising as polypharmacy is a known risk factor for mortality. Smoking behavior, number of medication and income level are ascertainable in patient consultation without requiring additional resources and may therefore be taken into account in health care measures.

Our analysis has several limitations. With a mean age of 56 years, the study population of the UKB is still relatively young, and accordingly there is a relatively low occurrence of death. This may have affected the power of our analysis. However, age-stratified analysis showed only slight differences between age groups. Secondly, a healthy volunteer bias has been observed for participants of the UK Biobank, with participants engaging less in harmful behavior, such as smoking and alcohol consumption, than the general population while living in less socioeconomically deprived areas (39). Future research may therefore assess the relevance of predictors in more marginalized populations. Further, data on lifestyle behavior in this analysis is self-reported, which comes with a risk of social desirability bias due to underreporting.

The strength of our analysis lies predominantly in the large sample size of the UK Biobank allowing for robust observations and exploration of relative contribution of clustering factors influencing mortality risk. Despite potentially limited representativeness, it has been suggested, that exposure-disease relationships are still extrapolatable to the general population (39). Further, the extensiveness of the UK Biobank database allowed us to adjust our analysis for a variety of lifestyle and health related factors. Additionally, the Oxford WebQ that was used to collect dietary data has been validated against interviewer-administered diet recalls and shown to produce reliable results (23, 40). Metabolites measured through the Nightingale platform have been applied in various epidemiologic studies and validated (41, 42).

## 5. Conclusion

We established associations of health- and lifestyle related factors with MetaboHealth, a molecular marker for global health and mortality. Among others, smoking, low income and an unhealthful plant-based were associated with higher adverse MetaboHealth. We further show that a predictor integrating the MetaboHealth score and especially smoking, income and medication intake improves mortality prediction beyond either the molecular of environmental factors only and especially for older adults and men. These findings may be applied when applying health checks and lifestyle interventions for prevention strategies in population health to improve identification of at-risk groups.

## Data Availability

Data is available from https://www.ukbiobank.ac.uk/use-our-data/

## 7. Statements & Declarations

## Acknowledgements

This research has been conducted using the UK Biobank Resource under Application Number 78275. This work has received funding from the European Union’s Horizon 2020 research and innovation framework under the Marie Sklodowska-Curie grant agreement No. 860173. P.E. Slagboom, M. Beekman, and L.C.P.G.M de Groot have received funding from the Vitality Oriented Innovations for the Lifecourse of the Ageing Society (VOILA) Consortium [ZonMw 457001001].

## Conflict of Interest

All authors declare no conflict of interest.

## Author Contributions

All authors contributed to the study conception and design. Analysis were performed by Kerstin Schorr. The first draft of the manuscript was written by Kerstin Schorr and all authors commented on previous versions of the manuscript. All authors read and approved the final manuscript.

## Ethical approval

UK Biobank has approval from the North West Multi-centre Research Ethics Committee (MREC) as a Research Tissue Bank (RTB) approval (https://www.ukbiobank.ac.uk/learn-more-about-uk-biobank/about-us/ethics).

## Footnotes

1 https://biobank.ndph.ox.ac.uk/showcase/ukb/docs/DietWebQ.pdf

## Notes

### Competing Interest Statement

The authors have declared no competing interest.

